# The feasibility of convalescent plasma therapy in severe COVID- 19 patients: a pilot study

**DOI:** 10.1101/2020.03.16.20036145

**Authors:** Kai Duan, Bende Liu, Cesheng Li, Huajun Zhang, Ting Yu, Jieming Qu, Min Zhou, Li Chen, Shengli Meng, Yong Hu, Cheng Peng, Mingchao Yuan, Jinyan Huang, Zejun Wang, Jianhong Yu, Xiaoxiao Gao, Dan Wang, Xiaoqi Yu, Li Li, Jiayou Zhang, Xiao Wu, Bei Li, Yanping Xu, Wei Chen, Yan Peng, Yeqin Hu, Lianzhen Lin, Xuefei Liu, Shihe Huang, Zhijun Zhou, Lianghao Zhang, Yue Wang, Zhi Zhang, Kun Deng, Zhiwu Xia, Qin Gong, Wei Zhang, Xiaobei Zheng, Ying Liu, Huichuan Yang, Dongbo Zhou, Ding Yu, Jifeng Hou, Zhengli Shi, Chen Saijuan, Zhu Chen, Xinxin Zhang, Xiaoming Yang

**Author notes:** Correspondence: Xiaoming Yang, China National Biotec Group Company Limited, National Engineering Technology Research Center for Combined Vaccines, Wuhan 430207, China, **Email:**, Xinxin Zhang, Research Laboratory of Clinical Virology, Ruijin Hospital and Ruijin Hospital North, National Research Center for Translational Medicine (Shanghai), Shanghai Jiao Tong University School of Medicine, No.197 Ruijin 2^nd^ Road, Shanghai 200025, China, **Email:**, Zhu Chen, State Key Laboratory of Medical Genomics, Shanghai Institute of Hematology, National Research Center for Translational Medicine (Shanghai), Ruijin Hospital, Shanghai Jiao Tong University School of Medicine, No.197 Ruijin 2**Email:**. These authors contributed equally (Kai Duan, Bende Liu, Cesheng Li, Huajun Zhang, Ting Yu, Jieming Qu, Min Zhou, Li Chen).

## Abstract

Currently, there are no approved specific antiviral agents for 2019 novel coronavirus disease (COVID-19). In this study, ten severe patients confirmed by real-time viral RNA test were enrolled prospectively. One dose of 200 mL convalescent plasma (CP) derived from recently recovered donors with the neutralizing antibody titers above 1:640 was transfused to the patients as an addition to maximal supportive care and antiviral agents. The primary endpoint was the safety of CP transfusion. The second endpoints were the improvement of clinical symptoms and laboratory parameters within 3 days after CP transfusion. The median time from onset of illness to CP transfusion was 16.5 days. After CP transfusion, the level of neutralizing antibody increased rapidly up to 1:640 in five cases, while that of the other four cases maintained at a high level (1:640). The clinical symptoms were significantly improved along with increase of oxyhemoglobin saturation within 3 days. Several parameters tended to improve as compared to pre-transfusion, including increased lymphocyte counts (0.65×10^9^/L vs. 0.76×10^9^/L) and decreased C-reactive protein (55.98 mg/L vs. 18.13 mg/L). Radiological examinations showed varying degrees of absorption of lung lesionswithin 7 days. The viral load was undetectable after transfusion in seven patients who had previous viremia. No severe adverse effects were observed. This study showed CP therapy was welltolerated and could potentially improve the clinical outcomes through neutralizing viremia in severe COVID-19 cases. The optimal dose and time point, as well as the clinical benefit of CP therapy, needs further investigation in larger well-controlled trials.

**Significance Statement:** COVID-19 is currently a big threat to global health. However, no specific antiviral agents are available for its treatment. In this work, we explored the feasibility of convalescent plasma (CP) transfusion to rescue severe patients. The results from 10 severe adult cases showed that one dose (200 mL) of CP was welltolerated and could significantly increase or maintain the neutralizing antibodies at a high level, leading to disappearance of viremia in 7 days. Meanwhile, clinical symptoms and paraclinical criteria rapidly improved within 3 days. Radiological examination showed varying degrees of absorption of lung lesions within 7 days. These results indicate that CP can serve as a promising rescue option for severe COVID-19 while the randomized trial is warranted.

## Main Text

### Introduction

Since December 2019, a pneumonia associated with severe acute respiratory syndrome coronavirus 2 (SARS-CoV-2), named as 2019 novel coronavirus disease (COVID-19) by World Health Organization (WHO), emerged in Wuhan, China (1-3). The epidemic spread rapidly worldwidewithin three months and was characterized as a pandemic by WHO on March 11,2020.As of March 12, 2020, a total of80,980confirmed cases and 3,173deaths had been reported in China. Meanwhile, a total of 44,377 confirmed cases and 1,446 deaths was reported in other 108 countries or regions.Currently, there are no approved specific antiviral agents targeting the novel virus, while some drugs are still under investigation, including remdesivir and lopinavir/ritonavir (4, 5). Although remdesivirwas reported to possess potential antiviral effectin one COVID-19 patient from the U.S., randomized controlled trials of this drug are ongoing to determine its safety and efficacy (6). Moreover, the corticosteroid treatment for COVID-19 lung injury remains controversial, due to delayed clearance of viral infection and complications(7,8). Since the effective vaccine and specific antiviral medicines are unavailable, it is an urgent need to look for an alternative strategy for COVID-19 treatment, especially amongsevere patients.

Convalescent plasma (CP) therapy, a classic adaptiveimmunotherapy, has been applied to the prevention and treatment of many infectious diseases for more than one century. Over the past two decades,CP therapy was successfully used in the treatment of SARS, MERS, and 2009 H1N1 pandemic with satisfactory efficacy and safety (9-12). A meta-analysis from 32 studies of SARS coronavirus infection and severe influenza showed a statistically significant reduction in the pooled odds of mortality following CP therapy, compared with placebo or no therapy (odds ratio, 0.25; 95% confidence interval, 0.14-0.45) (13). However, the CP therapy was unable to significantly improve the survival in the Ebola virus disease, probably due to the absence of data of neutralizing antibodytitrationfor stratified analysis (14). Since the virological and clinical characteristics share similarity among SARS, MERS, and COVID-19 (15), CP therapy might be a promising treatment option for COVID-19 rescue(16). Patients who have recovered from COVID-19 with a high neutralizing antibody titermay bea valuable donor source of CP. Nevertheless, the potential clinical benefit and risk of convalescent blood products in COVID-19 remains uncertain. Hence, we performed this pilot study in three participated hospitals to explore the feasibility of CPtreatment in 10 severe COVID-19 patients.

## Results

### Neutralizing activity of CP against SARS-CoV-2

The neutralizing activity against SARS-CoV-2 was evaluated by classical plaque reduction test using a recently isolated viral strain (1). Among the first batch of CP samples from 40 recovered COVID-19 patients, 39 showed high antibody titers of at least 1:160 whereas only one had a antibody titer of 1:32. This result laid the basis for our pilot clinical trial using CP in severe patients.

### General characteristics of Patients in the trial

From January 23, 2020, to February 19, 2020, ten severe COVID-19 patients (six males and four females) were enrolled and received CP transfusion. The median age was 52.5 years (IQR, 45.0– 59.5 years) (Table 1). None of the patients had direct exposure to Huanan Seafood Wholesale Market. The median time from onset of symptoms to hospital admission and CP transfusion was 6 days (IQR, 2.5–8.5 days) and 16.5 days (IQR11.0–19.3 days), respectively. Three patients were affected by clustering infection. The most common symptoms at disease onset were fever (seven of ten patients), cough (eight cases), and shortness of breath (eight cases), while less common symptoms included sputum production (five cases), chest pain (two cases), diarrhea (two cases), nausea and vomiting (two cases), headache (one case), and sore throat (one case). Four patients had underlying chronic diseases, including cardiovascular and/or cerebrovascular diseases and essential hypertension. Nine patients received arbidolmonotherapy or combination therapy with remdesivir (in one case not included in the current clinical trial), or ribavirin,or peramivir, while one patient received ribavirin monotherapy(Table 2). Antibacterial or antifungal treatment was used when patients had co-infection. Six patients received intravenousmethylprednisolone (20 mg every 24 hrs).

**Table 1.** Clinical characteristics of patients receiving convalescent plasma transfusion.

| No. | Sex | Age | Clinical classification | Days of admission from symptom onset | Days of convalescent plasma therapy from symptom onset | Clustering infection | Principal symptoms | Comorbidity |
| --- | --- | --- | --- | --- | --- | --- | --- | --- |
| 1 | M | 46 | Severe | 8 | 11 | No | Fever, cough, sputum production, shortness of breath, chest pain | Hypertension |
| 2 | F | 34 | Severe | 0 | 11 | Yes | Cough, shortness of breath, chest pain, nausea and vomiting | None |
| 3 | M | 42 | Severe | 8 | 19 | Yes | Fever, cough, sputum production, shortness of breath, sore throat, diarrhea | Hypertension |
| 4 | F | 55 | Severe | 10 | 19 | No | Fever, cough, sputum production, shortness of breath | None |
| 5 | M | 57 | Severe | 4 | 14 | No | Fever, shortness of breath | None |
| 6 | F | 78 | Severe | 8 | 17 | Yes | Fever, cough, sputum production, shortness of breath, muscle ache | None |
| 7 | M | 56 | Severe | 4 | 16 | No | Fever, cough, sputum production, arthralgia | None |
| 8 | M | 67 | Severe | 10 | 20 | No | Fever, cough, headache, diarrhea, vomiting | Cardiovascular and cerebrovascular diseases |
| 9 | F | 49 | Severe | 1 | 10 | No | Cough, shortness of breath | None |
| 10 | M | 50 | Severe | 3 | 20 | No | Shortness of breath | Hypertension |

M=male. F=female.

**Table 2.** Other treatments of ten patients receiving convalescent plasma transfusion.

| No. | Drugs administered |  |  | Oxygen support |  |
| --- | --- | --- | --- | --- | --- |
|  | Antiviral treatment | Antibiotic or antifungal treatment | Corticosteroids treatment | Before convalescent plasma therapy | After convalescent plasma therapy |
| 1 | Arbidol0.2g q8h po.<br>Ribavirin 0.5g qdi.v. | Cefoperazone Sodium i.v. | None | High-flow nasal cannula,<br>mechanical ventilation | Mechanical ventilation |
| 2 | Arbidol0.2g q8hpo. | Cefoperazone Sodium i.v. | None | None | None |
| 3 | Arbidol0.2g q8hpo. | Moxifloxacin i.v. | Methylprednisolone i.v. | High-flow nasal cannula,<br>mechanical ventilation | High-flow nasal cannula |
| 4 | Ribavirin 0.5g qdi.v. | Linezolid i.v.Imipenem -<br>Sitastatin Sodium i.v. | Methylprednisolone i.v. | Mechanical ventilation | High-flow nasal cannula |
| 5 | Arbidol0.2g q8hpo.<br>Remdesivir0.2gqdi.v.<br>Interferon-α<br>500MIUqdinh. | Moxifloxacin i.v.Cefoperagone Sodium<br>and Tazobactam Sodium i.v. | Methylprednisolone i.v. | Low-flow nasal cannula | Low-flow nasal cannula |
| 6 | Arbidol0.2g q8h po. | Cefoperazone Sodium i.v.<br>Levofloxacin i.v. | Methylprednisolone i.v. | High-flow nasal cannula | High-flow nasal cannula |
| 7 | Arbidol0.2g q8h po. | Cefoperagone Sodium and<br>Tazobactam Sodium i.v.<br>Fluconazole i.v. | Methylprednisolone i.v. | High-flow nasal cannula | none |
| 8 | Arbidol0.2g q8h po.<br>Ribavirin 0.5g qdi.v. | None | None | None | None |
| 9 | Arbidol0.2g q8h po.<br>Oseltamivir75mg<br>q12h po.<br>Peramivir0.3gqdi.v. | None | None | Low-flow nasal cannula | Low-flow nasal cannula<br>(Intermittent) |
| 10 | Arbidol0.2g q8h po.<br>Interferon-α 500MIU<br>qdinh. | Cefoperazone Sodium<br>i.v.Casprofungini.v. | Methylprednisolone i.v. | High-flow nasal cannula | High-flow nasal cannula |

po.=peros. i.v.=intravenous injection. inh.=inhalation.

On computer-assisted tomography (CT), all patients presented bilateral ground-glass opacity and/or pulmonary parenchymal consolidation with predominantly subpleural and bronchovascular bundles distribution in the lungs. Seven patients had multiple lobe involvement and four patients had interlobular septal thickening.

### Effects of CP transfusion

#### Improvement of clinical symptoms

All symptoms inthe 10 patients, especially fever, cough, shortness of breath and chest pain, disappeared or largely improved within 1-3 days upon CP transfusion. Prior toCP treatment, three patients received mechanical ventilation, three received high-flow nasal cannula oxygenation, and two received conventional low-flow nasal cannula oxygenation. After treatmentwith CP, two patients were weaned from mechanical ventilation to high-flow nasal cannula and one patient discontinued high flow nasal cannula. Besides, in one patient treated with conventional nasal cannula oxygenation, continuous oxygenation was shifted to intermittent one (Table 2).

#### Reduction of pulmonary lesions on chest CT examinations

According to chest CTs, all patients showed different degrees of absorption of pulmonary lesions after CP transfusion. Representative chest CT images of patient 9 and patient 10 were shown on Fig. 1. Patient 9, a 49-year-old female admitted on 1 day post onset of illness (dpoi), showed the most obvious pulmonary image improvement. On 10 dpoi, one dose of 200 mL transfusion of CP was given. The SARS-CoV-2 RNA converted to negative on 12 dpoi. Compared with the result on 7 dpoi, massive infiltration and ground-glass attenuation disappeared on CT image performed on 13 dpoi, accompanied bya much better pulmonary function. Patient 10, a 50-year-old male, was admitted on 3 dpoi and was given a 200 mL transfusion of CP on 20 dpoi. His chest CT presented massive infiltration and widespread ground-glass attenuation on admission andstarted to show a gradual absorption of lung lesions 5 days after CP transfusion. The SARS-CoV-2 RNA became negative on 25 dpoi.

**Figure 1.**
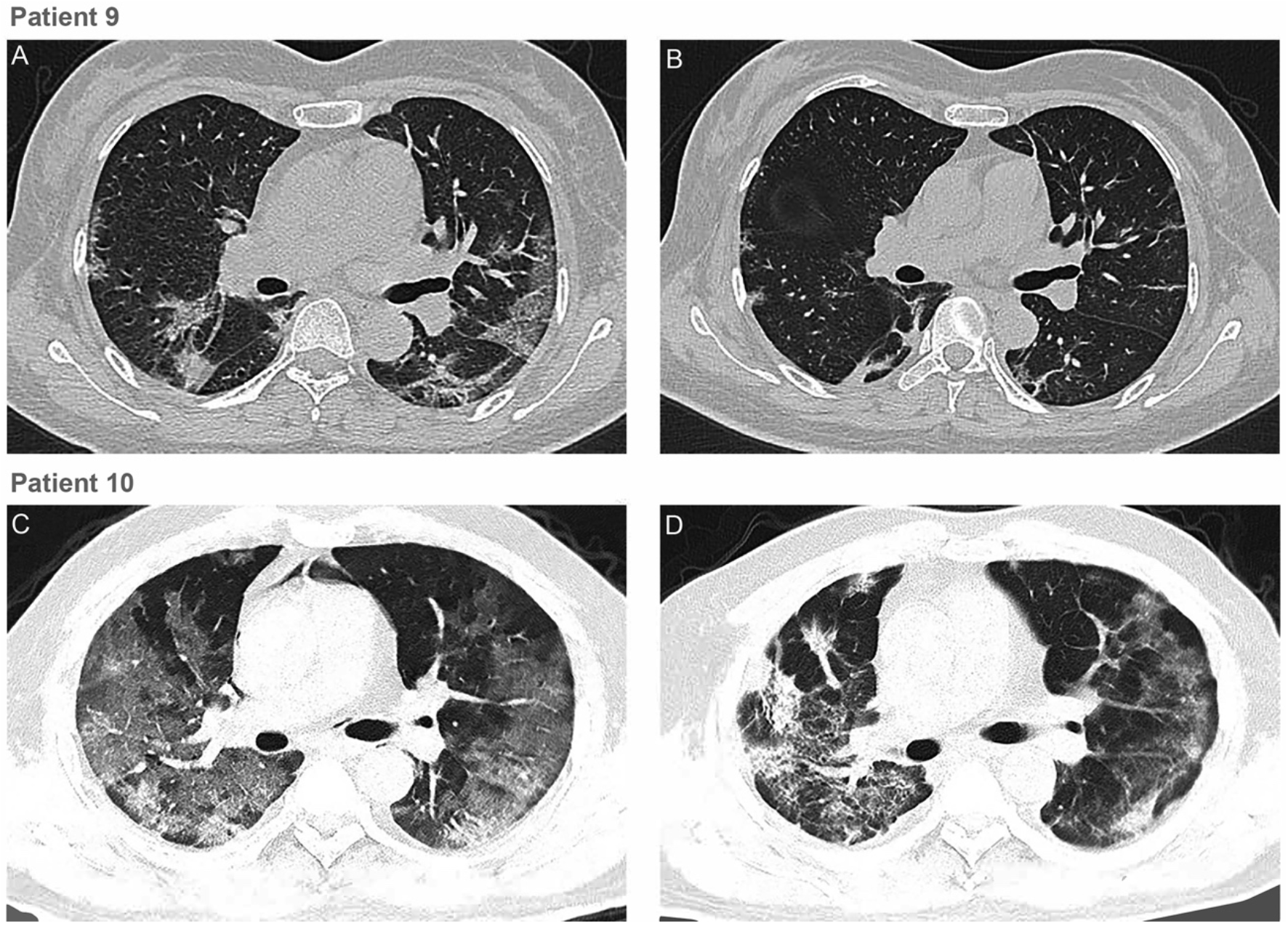
Chest CTs of two patients (*A*) Chest CT of patient 9 obtained on Feb 9 (7 dpoi) before convalescent plasma transfusion (10 dpoi)showed ground glass opacity with uneven density involving the multilobal segments of both lungs. The heart shadow outline was not clear. The lesion was close to the pleura. (*B*) CT Image of patient 9 taken on Feb 15(13 dpoi) showed the absorption of bilateral ground glassopacity after convalescent plasma transfusion. (*C*)Chest CT of patient 10 was obtained on Feb 8 (19 dpoi) before convalescent plasma transfusion (20 dpoi). The brightness of both lungs was diffusely decreased and multiple shadows of high density in both lungs were observed.(*D*) Chest CTof patient 10 on Feb 18 (29dpoi) showed those lesions improvedafter convalescent plasma transfusion.

#### Amelioration ofroutine laboratory criteria and pulmonary function

Lymphocytopenia, an important index for prognosis inCOVID-19 (2), tended to beimprovedafter CP transfusion (median: 0.65×10^9^ per L vs. 0.76×10^9^ per L), seven out of ten patients showingan increase of lymphocyte counts (Fig. 2). Concerning other laboratory tests, we observed a tendency of decrement of parameters indicative of inflammation and/or liver dysfunction as compared to the statusbefore CP therapy. These included C-reactive protein (CRP) (median: 55.98 mg/L vs. 18.13 mg/L), alanine aminotransferase (median: 42.00 U/L vs. 34.30 U/L) and aspartate aminotransferase (median: 38.10 U/L vs. 30.30 U/L) (Table 3). The total bilirubin (median: 12.40 μmol/L vs. 13.98 μmol/L) remained unchanged except an obvious increment in patient 1 (Fig. 2). An increase of SaO_2_ (median:93.00% vs. 96.00%), a measurement constantly performed in most patients in our trial, was found, which could indicaterecovering lung function.This temporal relationship was notable despite the provision of maximal supportive care and antiviral agents.

**Table 3.** Comparison of laboratory parameters before and after convalescent plasma transfusion

| Clinical Factors | Before CP transfusion | After CP transfusion |
| --- | --- | --- |
| C-reactive protein (mg/L, normal range 0-6) | 55.98<br>(15.57-66.67) | 18.13<br>(10.92-71.44) |
| Lymphocyte ($10^9$ per L, normal range 1.1-3.2) | 0.65<br>(0.53-0.90) | 0.76<br>(0.52-1.43) |
| Alanine aminotransferase (U/L, normal range 9-50) | 42.00<br>(28.25-61.85) | 34.30<br>(25.75-53.90) |
| Aspartate aminotransferase (U/L, normal range 15-40) | 38.10<br>(28.50-44.00) | 30.30<br>(17.30-38.10) |
| Total bilirubin (μmol/L, normal range 0-26) | 12.40<br>(11.71-22.05) | 13.98<br>(12.20-20.80) |
| SaO<sub>2</sub> (% , normal range $\geq 95$) | 93.00<br>(89.00-96.50) | 96.00<br>(95.00-96.50) |

**Figure 2.**
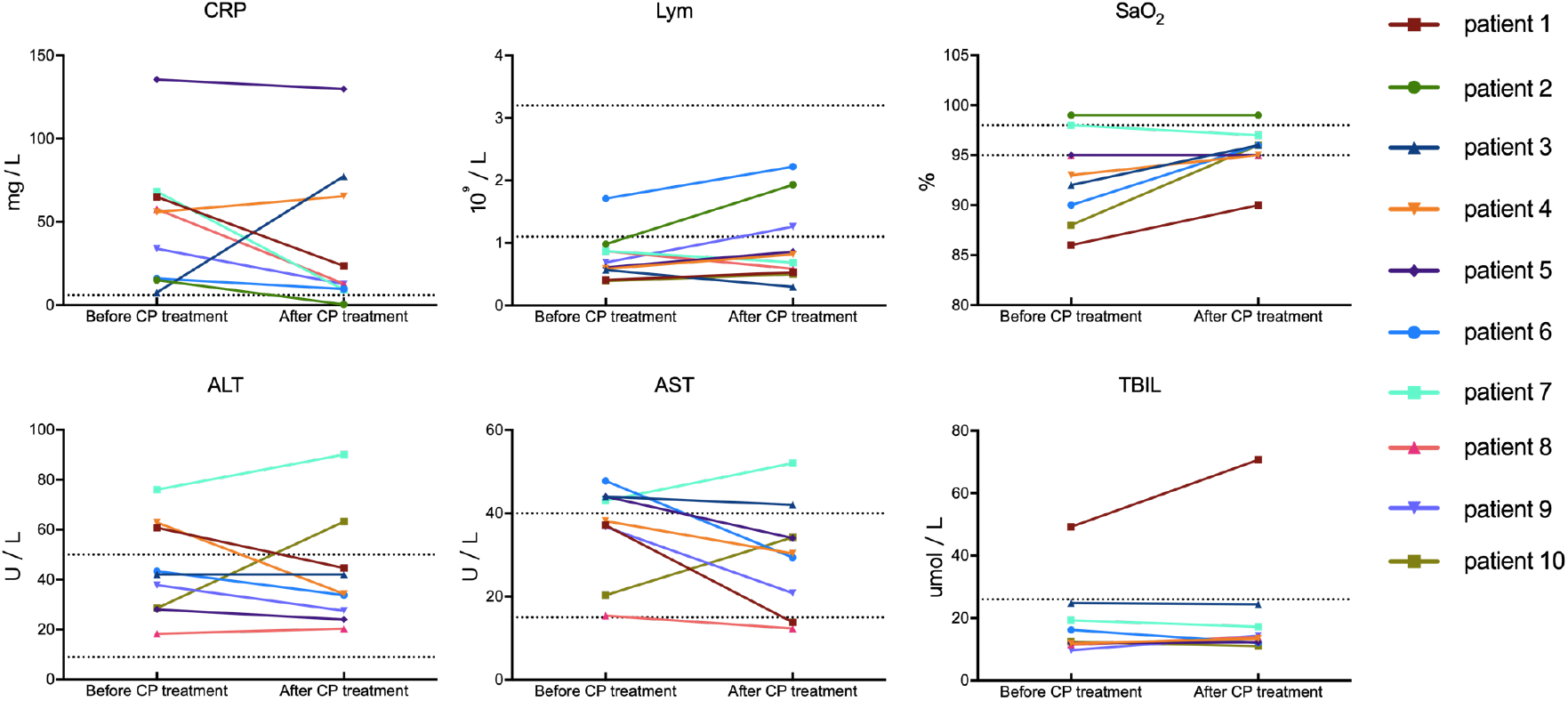
Dynamic changes of laboratory parameters in all patients. The dotted horizontal line represents the reference value range. CP=convalescent plasma. CRP=C-reactive protein. SaO_2_=oxyhemoglobin saturation. TBIL=total bilirubin. ALT=alanine aminotransferase. AST=aspartate aminotransferase.

Remarkably, patient 1, a 46-year-old male admitted on 8 dpoi, had a very quick recovery with much improved result of laboratory tests. He received antiviral drugs (arbidol and ribavirin) treatment and high flow nasal cannula on admission. Mechanical ventilation was given on 10 dpoi for critical care support. CP transfusion was performed on 11 dpoi. On 12 dpoi, the SARS-CoV-2test turned to negative, with a sharp decrease of CRP from 65.04 mg/L to 23.57 mg/L and incrementof SaO_2_ from 86% to 90% (Fig. 3). The mechanical ventilation was successfully weaned off 2 days after CP transfusion. On 15 dpoi, a steady elevation oflymphocyte count and a drop ofaminopherase levelwere observed, indicating improvement ofimmunological and hepatic function.

**Figure 3.**
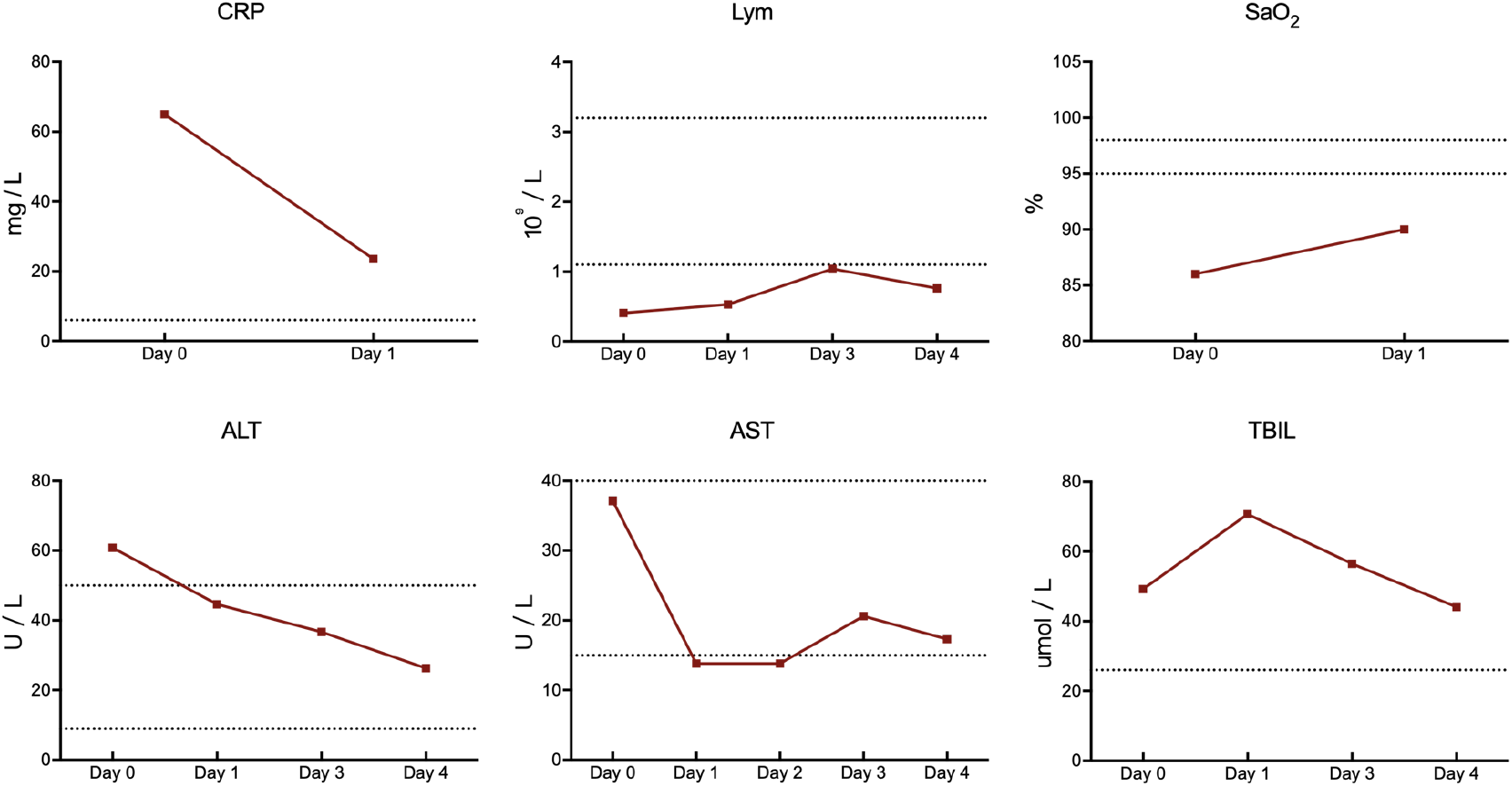
Change of laboratory parameters in patient 1 X-axis represents the day post convalescent plasma transfusion. The dotted horizontal line represents the reference value range. CP=convalescent plasma. CRP=C-reactive protein. SaO_2_=oxyhemoglobin saturation. TBIL=total bilirubin. ALT=alanine aminotransferase. AST=aspartate aminotransferase.

#### Increase of neutralizing antibody titers and disappearanceof SARS-CoV-2 RNA

We determined neutralizing antibody titers before and after CP transfusion in all patients except one (patient2) (Table 4). The neutralizing antibody titers of five patients increased and four patients remained atthe same level after CP transfusion.SARS-CoV-2 RNA, assayed by reverse transcriptase-polymerase chain reaction (RT-PCR), was positive in seven patients and negative in three cases before CP transfusion. Of note, SARS-CoV-2 RNA was decreased to anundetectable level in 3 patients on day 2, 3 patients on day 3 and 1 patients on day 6 after CP therapy.These results were in support of anneutralizing effectof CP on serum SARS-CoV-2.

**Table 4.** Comparison of serum neutralizing antibody titers and SARS-CoV-2 RNA load before and after convalescent plasma therapy

| patient No. | CP transfusion Date | Before CP transfusion |  |  | After CP transfusion |  |  |
| --- | --- | --- | --- | --- | --- | --- | --- |
|  |  | Date | Serum neutralizing antibody titres | Serum SARS-CoV-2 RNA load (Ct value) | Date | Serum neutralizing antibody titres | Serum SARS-CoV-2 RNA load (Ct value) |
| 1 | February 9 | February 8 | 1:160 | 37.25 | February 10 | 1:640 | negative |
| 2 | February 9 | February 8 | Unavailable | 35.08 | February 11 | Unavailable | negative |
| 3 | February 13 | February 12 | 1:320 | 38.07 | February 14 | 1:640 | negative |
| 4 | February 13 | February 12 | 1:160 | 37.68 | February 14 | 1:640 | negative |
| 5 | February 12 | February 11 | 1:640 | negative | February 14 | 1:640 | negative |
| 6 | February 12 | February 11 | 1:640 | negative | February 14 | 1:640 | negative |
| 7 | February 12 | February 11 | 1:320 | 34.64 | February 14 | 1:640 | negative |
| 8 | February 12 | February 11 | 1:640 | 35.45 | February 14 | 1:640 | negative |
| 9 | February 12 | February 11 | 1:160 | negative | February 14 | 1:640 | negative |
| 10 | February 9 | February 8 | 1:640 | 38.19 | February 14 | 1:640 | negative |

#### Outcome of patients treated with CP as compared to a recent historic control group

Ahistoric control group was formed by random selection of10 patients from the cohort treated in the same hospitals andmatched by age, gender and severity of the diseases to the 10 cases in our trial. Baseline characteristics of patients between CP treatment group and control group showed no significant differences, while clinical outcomesof these two groups weredifferent: 3 cases dischargedwhile 7 cases in much improved status and ready for discharge in CP group, as compared to 3 deaths, 6 cases in stablized status and one case in improvement in the control group (p<0.001, Supplementary table 1).

#### Adverse effects of CP transfusions

Patient 2 showed an evanescent facial red spot. No serious adverse reactions or safety events were recorded after CP transfusion.

## Discussion

To our knowledge, this is the first study to explore the feasibility of CP therapy in COVID-19. All enrolled severe COVID-19 patients achieved primary and secondary outcomes. One dose of 200 mL CP transfusion was welltolerated, while the clinical symptoms significantly improved with the increase of oxyhemoglobin saturation within 3 days, accompanied by rapid neutralization of viremia. Severe pneumonia caused by human coronavirus was characterized by rapid viral replication, massive inflammatory cell infiltration, and elevated proinflammatory cytokines or even cytokine storm in alveoli of lungs, resulting in acute pulmonary injury and acute respiratory distress syndrome (ARDS) (17). Recent studies on COVID-19 demonstrated that the lymphocyte counts in the peripheral blood were remarkably decreased and the levels of cytokines in the plasma from patients requiring ICU support, including IL-6, IL-10, TNF-ɑ, GM-CSF, were significantly higher than those who did not require ICU conditions (2, 18). CP, obtained from recovered COVID-19 patients who had established humoral immunity against the virus, contains a large quantity of neutralizing antibodies capable of neutralizing SARS-CoV-2 and eradicating the pathogen from blood circulation and pulmonary tissues (19). In the present study, all investigated patients achieved serum SARS-CoV-2 RNA negativity after CP transfusion, accompanied by the increase of oxygen saturation and lymphocyte counts, and the improvement of liver function and C-reactive protein. The results suggested that the inflammation and overreaction of the immune system were alleviated by antibodies contained in CP. The case-fatality rates (CFRs) in the present study were 0% (0/10), which was comparable to the CFRs in SARS which varied from 0% (0/10) to 12.5% (10/80) in four non-comparative studies using CP treatment (9, 20-22). Based on our preliminary results, CP therapy can be an easy-accessible, promising and safe rescue option for severe COVID-19 patients. It is nevertheless worth mentioning that the absorption of pulmonary lesions was often behind the improvement of clinical symptoms, as shown in patients 9 and 10 in this trial. The first key factor associated with CP therapy is the neutralizing antibody titer. A small sample study in MERS-CoV infection showed that the neutralizing antibody titer should exceed 1:80 to achieve effective CP therapy (12). To find eligible donors who have high levels of neutralizing antibody is a prerequisite. Cao (23) et al showed that the level of specific neutralizing antibody to SARS-CoV decreased gradually 4 months after the disease process, reaching undetectable levels in 25.6% (IgG) and 16.1% (neutralizing antibodies) of patients at 36 months after disease status. A study from the MERS-CoV infected patients and the exposed healthcare workers showed that the prevalence of MERS-CoVIgGseroreactivity was very low (2.7%), and the antibodies titer decreased rapidly within 3 months (24). These studies suggested that the neutralizing antibodies represented short-lasting humoral immune response and plasma from recently recovered patients should be more effective. In the present study, recently recovered COVID-19 patients, who were infected by SARS-CoV-2 with neutralizing antibody titer above 1:640and recruited from local hospitals should be considered as suitable donors. The median age of donors was lower than that of recipients (42.0 vs. 52.5 years). Among the nine cases investigated, the neutralizing antibody titers of five patients increased while four patients kept the same level to 1:640 within two days. The antibody titers in CP in COVID-19 seem thus higher than those used in the treatment of MERS patient (1:80) (12). The second key factor associated with efficacy is the treatment time point. A better treatment outcome was observed among SARS patients who were given CP before 14 dpoi (58.3% vs 15.6%; *P* < 0.01), highlighting the importance of timely rescue therapy (9). The mean time from onset of illness to CP transfusion was 16.5 days. Consistent with previous research, all three patients receiving plasma transfusion given before 14 dpoi (patients 1, 2 and 9) in our study showed a rapid increase of lymphocyte counts and a decrease of CRP, with remarkable absorption of lung lesions in CT. Notably, patients who received CP transfusion after 14 dpoi showed much less significant improvement, such as patient 10. However, the dynamics of the viremia of SARS-CoV-2 was unclear, so the optimal transfusion time point needs to be determined in the future.

In the present study, no severe adverse effects were observed. One of the risks of plasma transfusion is the transmission of the potential pathogen. Methylene blue photochemistry was applied in this study to inactivate the potential residual virus and to maintain the activity of neutralizing antibodies as much as possible, a method known to be much better than ultraviolet C light (25). No specific virus was detected before transfusion. Transfusion-related acute lung injury (TRALI) was reported in an Ebola virus disease woman who received CP therapy (26). Although uncommon in the general population receiving plasma transfusion, this specific adverse reaction is worth noting, especially among critically illed patients experiencing significant pulmonary injury (27). Another rare risk worth mentioning during CP therapy is antibody-dependent infection enhancement, occurring at sub-neutralizing concentrations, which could suppress innate antiviral systems and thus could allow logarithmic intracellular growth of the virus (28). The special immune enhancement was reportedly more common in Dengue fever, but also could be found in SARS-CoV infection *in vitro* (29). No such pulmonary injury and infection enhancement were observed in our patients, probably owing to high levels of neutralizing antibodies, timely transfusion, and appropriate plasma volume.

There were some limitations to the present study. First, except for CP transfusion, the patients received other standard cares. All patients received antiviral treatment despite the uncertainty of the efficacy of drugs used. As a result, the possibility thatthese antiviral agents could contribute to the recovery of patients, or synergize with the therapeutic effect of CP, could not be ruled out. Furthermore, some patients received glucocorticoid therapy, which might interfere with immune response and delay virus clearance. Second, the median time from onset of symptoms to CP transfusion was 16.5 days (IQR11.0-19.3 days). Although the kinetics of viremia during natural history remains unclear, the relationship between SARS-CoV-2 RNA reduction and CP therapy, as well asthe optimal concentration of neutralizing antibodies and treatment schedule, should be further clarified. Third, the dynamic changes of cytokines during treatment were not investigated. Nevertheless, the preliminary results of this trial seem promising, justifying a randomized controlled clinical trial in a larger patient cohort.

In conclusion, this pilot study on CP therapy showed a potential therapeutic effect and low risk in the treatment of severe COVID-19 patients. One dose of CP with high concentration of neutralizing antibodies can rapidly reduce the viral load and tends to improve clinical outcomes. The optimal dose and treatment time point, as well as the definite clinical benefits of CP therapy, need to be further investigated in randomized clinical studies.

## Materials and Methods

### Patients

From January 23, 2020, to February 19, 2020, ten patients in three participating hospitals (Wuhan Jinyintan Hospital, the Jiangxia District Hospital of Integrative Traditional Chinese and Western Medicine, Wuhan, and the First People’s Hospital of Jiangxia District, Wuhan) were recruited in this pilot study. All patients were diagnosed as having severe COVID-19 according to the WHO Interim Guidance (30) and the Guideline of Diagnosis and Treatment of COVID-19 of National Health Commission of China (version 5.0) (31), with confirmation by real-time RT-PCR assay. The enrollment criteria were one of the conditions (2 to 4) plus condition (1): 1). Age ≥18 years; 2).Respiratory distress, RR ≥30 beats/min; 3).Oxygen saturation level less than 93% in resting state; 4). Partial pressure of oxygen (PaO_2_)/oxygen concentration (FiO_2_) ≤300 mmHg (1 mmHg=0.133 kPa). The exclusion criteria were as follows: 1). Previous allergic history to plasma or ingredients (Sodium Citrate); 2). Cases with serious general conditions, such as severe organ dysfunction, who were not suitable for CP transfusion; Written informed consent according to the Declaration of Helsinki was obtained from each patient or legal relatives. This study was approved by the Ethics Committee of the China National Biotec Group Co., Ltd. (Approval number:2020-0001). The registration number of this trial was ChiCTR2000030048.

### Donors for convalescent plasma transfusion

Tendonor patients who recovered from COVID-19 were recruited from three participating hospitals. The recovery criteria were as follows: 1). Normality of body temperature for more than 3 days; 2). Resolution of respiratory tract symptoms; 3). Two consecutively negative results of sputum SARS-CoV-2 of real-time reverse transcriptase-polymerase chain reaction (RT-PCR) assay (one-day sampling interval). The donor’s blood was collected after three weeks post-onset of illness and 4 days post-discharge. Written informed consent was obtained from each patient.

### Plasma preparation procedure and quality control

Apheresis was performed using a Baxter CS 300 cell separator (Baxter, Deerfield, IL, USA). Convalescence plasma for treatment was collected from 40donors. The median age was 42.0 years (IQR, 32.5–49 years). A 400–600 mL ABO-compatible plasma sample was harvested from each donor depending on the age and body weight, and each sample was divided and stored as 200 mL aliquots at 42103 without any detergent or heat treatment. The CP was then treated with methylene blue and light treatment for 30 minutes in the medical plasma virus inactivation cabinet (Shandong Zhongbaokang Medical Appliance Co., Ltd).

### Serology test and real-time RT–PCR detection of SARS-CoV-2 and other pathogens

The neutralized activity of plasma was determined by plaque reduction neutralization test using SARS-CoV-2 virusin the high biosafy level (BSL-4) laboratory of Wuhan Institute of Virology, Chinese Academy of Sciences.Neutralization titer was defined as the highest serum dilution with 50 % reduction in the number of plaques, as compared with the number of plaques in wells in the absence of novel coronavirus antibody as blank control. The neutralization activity of the receptor-binding domain (RBD) of antibody in the CP was detected by a sandwich ELISA.SARS-CoV-2-IgG antibody titer was tested by enzyme-linked immunosorbent assay. SARS-CoV-2 RNA was detected by RT-PCR assay and the result was presented as cycle threshold (Ct) value (Shanghai BioGerm Medical Biotechnology Co., Ltd). Methylene blue residue was detected by the verified ultraviolet method. The serology screening for hepatitis B and C virus, human immunodeficiency virus, and syphilis spirochete was negative. The protocols for SARS-CoV-2 serology and RNAtest are presented in the supplementary materials.

### Treatment

All patients were admitted to the intensive care unit (ICU) and received antiviral therapy and other supportive care, while somepatients received antibiotic treatment, antifungal treatment, glucocorticoid and oxygen support at the appropriate situation. One dose of 200 mL inactivated CP with neutralization activity >1:640 was transfused into the patients within 4 hours following the WHO blood transfusion protocol.

### Data collection

Clinical information of all enrolled patients was retrieved from the hospital electronic history system, including the baseline demographic data, days of illness duration, presenting symptoms, different kinds of examination and methods of treatment. Bacterial co-infection was identified by a positive culture from respiratory, urinary or blood culture within 48h of hospital admission. Complications including acute renal failure, acute coronary syndrome, myocarditis, acute respiratory distress syndrome, and nosocomial infection were recorded. The applications of assisted mechanical ventilation, intranasal oxygen inhalation, and medication regimen were recorded. The SARS-CoV-2 RNA from the serum sample was monitored during treatment.

### Outcome Measures and Definitions

The clinical symptoms were recorded by attending physicians daily. The blood test and biochemical tests were carried out every 1-2 days. SARS-CoV-2 RNA was detected every 2-3 days. CT scan was repeated every 3-5 days. The primary endpoint was the safety of CP transfusion. The second endpoints were the improvement of clinical symptoms, laboratory and radiological parameters within 3 days after CP transfusion. The clinical symptoms improvement was defined as temperature normalization, relief of dyspnea, and oxygen saturation normalization, and the radiological improvement was defined as different degrees of absorption of lung lesions.

### Statistical analysis

Continuous variables were presented as the median and interquartile range (IQR). Graphs were plotted using GraphPad Prism 7.0. Statistical software used included SPSS 24.0.

## Data Availability

All data associated with the manuscript were available on request

## Data Availability statement

The data that support the findings of this study are available from the corresponding author on reasonable request. Participant data without names and identifiers will be made available after approval from the corresponding author. After publication of study findings, the data will be available for others to request. The research team will provide an email address for communication once the data are approved to be shared with others. The proposal with detailed description of study objectives and statistical analysis plan will be needed for evaluation of the reasonability to request for our data. The corresponding author will make decision based on these materials. Additional materials may also be required during the process.

## Author Contributions

XZ, ZC, and XY contributed to the design of the study. SM, ZW, LL, JZ, WC, YH, SH, LZ, ZZ, ZX, JH, HY, DZ, and DY collected the epidemiological and clinical data. JH, XY, YX, XL, and JZ processed statistical data. KD, BL, CL, HZ, TY, JQ, MZ, ZC and LC drafted the manuscript. ZS, CP, XG, BL, YH, JY, XW, YP, LL, ZZ, YW, KD, QG, WZ, XZ, YL, MY, SC, and DW was responsible for virus detection and summarizing all epidemiological and clinical data. All authors reviewed and approved the final version.

## Acknowledgments

This study was funded by Key projects of the Ministry of Science and Technology China “Preparation of specific plasma and specific globulin from patients with a recovery period of COVID-19 infection” (project number: 2020YFC0841800).This work was also supported by Shanghai Guangci Translational Medicine Development Foundation. We thank all patients and donors involved in this study.

